# Red Blood Cell-Derived Exosomal Oncogenic MicroRNA Promote Cancer Development and Progression

**DOI:** 10.1101/2024.05.10.24307177

**Authors:** Jin Li, Pushpa Dhilipkannah, Van K Holden, Ashutosh Sachdeva, Feng Jiang

## Abstract

The role of red blood cells (RBCs) in tumorigenesis is poorly understood. We previously identified RBC-microRNAs with aberrations linked to lung cancer, including miR-93-5p. Here we find that miR-93-5p levels are elevated in RBC-derived exosomes among lung cancer patients and are associated with their shorter survivals. RBC-derived miR-93-5p transfers to cancer cells primarily through the exosomal pathway. The transferred RBC-miR-93-5p can target PTEN in cancer cells, and hence increase cell proliferation, invasion, and migration. RBC-derived miR-93-5p accelerates, whereas targeting miR-93-5p diminishes tumor growth in xenograft models. These findings reveal a novel biological function of RBCs in tumorigenesis, where they facilitate cancer progression by transferring the oncomiR via exosomes, thereby offering new diagnostic and treatment strategies for lung cancer.

## Introduction

Red blood cells (RBCs), conventionally recognized for their essential role in oxygen transport and regulation of the body’s metabolic demands. However, RBCs have recently been uncovered to play multifaceted roles beyond their conventional functions, including engaging in phagocytosis, antimicrobial defense, antigen recognition, and immune adherence (*1, 2*). Furthermore, an association between RBC and cancer has been observed (*2*). For example, RBCs can be present in the tumor microenvironment due to the leaky and disorganized vasculature commonly associated malignant lesions (*3, 4*). Hypoxia within tumors, a condition caused by inadequate oxygen supply by RBCs, can lead to the selection of more aggressive cancer cell phenotypes, and increase resistance to chemotherapy and radiation therapy(*5*). Liang et al. recently demonstrated that mature RBCs can acquire DNA from lung cancer tissues, and the acquired mutated DNA could be used for early cancer detection (*1*). However, the contribution of RBCs to tumorigenesis and the mechanisms behind this remain unclear.

Exosomes, which range in size from 20 to 200 nanometers, are essential in cellular communication by transporting DNA, various types of RNA, and protein fragments from their cell of origin to recipient cells (*6*). Furthermore, exosomes are integral to numerous physiological and pathological processes, particularly in the context of cancer progression and metastasis (*7*). For instance, exosomes can alter the tumor microenvironment by facilitating angiogenesis in eluding the immune response (*7*). Additionally, exosomes can modulate the behavior of distant cells and facilitate the preparation of metastatic niches by transferring a diverse array of molecules (*7*).

microRNAs (miRNAs) play a crucial role in regulating gene expression at the post-transcriptional level. miRNAs can function as oncomiRs or tumor suppressors by influencing key pathways involved in cell growth, cell death, and spread, thereby driving tumorigenesis (*8*). Furthermore, miRNAs within exosomes derived from cancer cells are pivotal in regulating processes related to malignancy (*9*). By transferring miRNAs between cells, exosomes can modulate gene expression in recipient cells, affecting cell proliferation, survival, and migration(*10*). Additionally, exosomal miRNAs from nucleated cells like fibroblasts, endothelial cells, mesenchymal stem cells, and immune cells can suppress tumor suppressors or enhance oncogenes, promoting tumor growth and chemotherapy resistance (*11*). However, due to the absence of a nucleus and associated machinery in RBCs, their derived miRNAs were previously considered to have minimal biological impact (*12*). Interestingly, we recently discovered a distinctive miRNA profile in RBCs, featuring miR-93-5p, which is closely associated with lung cancer (*13*). Notably, miR-93-5p is recognized as an oncomiRNA that crucially impacts cancer progression (*14, 15*). Our current study aims to elucidate the role of RBCs in tumorigenesis, particularly in lung cancer. We have discovered a novel biological role for RBCs in tumor progression, where they facilitate the transfer of oncogenic microRNAs via exosomal pathways.

## Results

### Elevated levels of miR-93-5p in RBCs and their derived exosomes are associated with poorer outcomes in lung cancer patients

To investigate the association of RBC-miR-93-5p with clinical outcomes, we quantified miR-93-5p and miR-451 (the housekeeper miRNA of RBCs) in RBCs, their corresponding exosomes and plasma samples from 58 lung cancer patients and 45 cancer-free smokers (table S1). In patients with lung cancer, miR-93-5p levels were significantly higher in both RBCs and their exosomes when compared to controls (all p<0.01) (Fig. 1A). However, this miRNA was not detectable in the plasma specimens of both cases and controls (Fig. 1A). A strong correlation was found between miR-93-5p levels in RBCs and their exosomes specifically in lung cancer patients (r=0.732, p=0.001) (Fig. 1B). Furthermore, higher levels of miR-93-5p in RBCs and exosomes were associated with lower survival rates, a history of smoking, and advanced stages of lung cancer (all p<0.05) (Fig. 1C) (table S2). miR-451 is detectable across all examined sample types; however, there are no significant differences in its levels between patient and control groups (all p>0.05) (Fig. 1D).

**Fig. 1.**
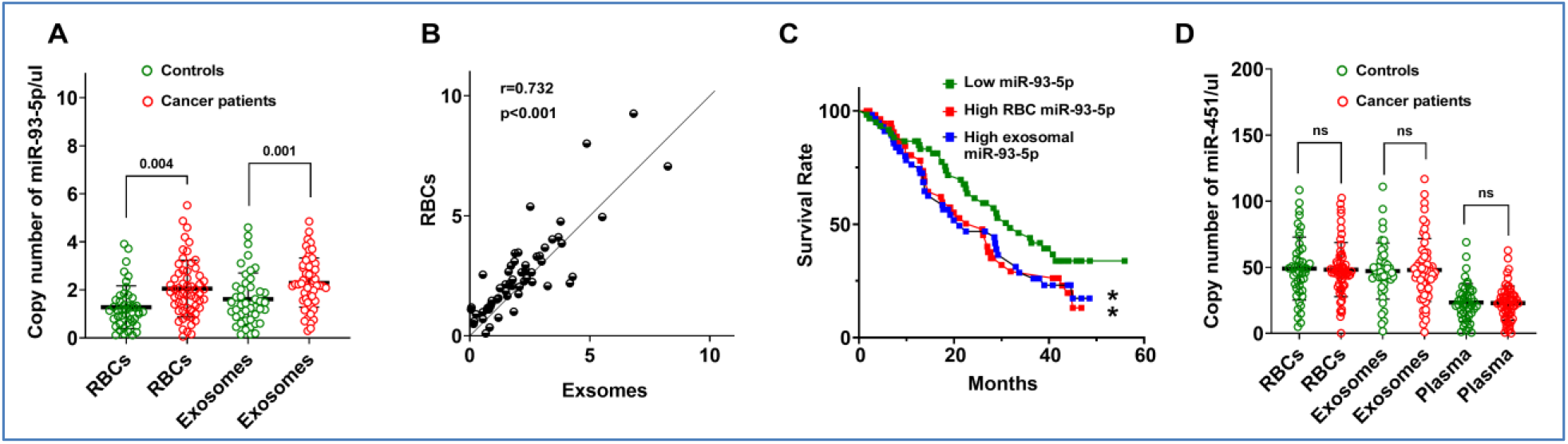
Analysis of miR-93-5p and miR-451 in lung cancer patients and cancer-free smokers (controls). (A) The expression of miR-93-5p is significantly higher in the RBCs and RBC-derived exosomes of lung cancer patients compared to controls (all p≤ 0.01, t-test). The expression of miR-93-5p is undetectable in plasma for both groups. (B) The correlation plot displays a positive relationship between miR-93-5p levels in RBCs and their derived exosomes in lung cancer patients, with a Pearson’s correlation coefficient of r=0.732 and p<0.01. (C) Kaplan-Meier survival curves for lung cancer patients categorized by levels of miR-93-5p in their RBCs and exosomes. The green line represents patients with low levels of miR-93-5p in both RBCs and exosomes, the red line corresponds to patients with high levels of miR-93-5p in RBCs, and the blue line is for patients with high levels of miR-93-5p in exosomes. Patients with higher miR-93-5p levels in RBCs or exosomes have reduced survival rates compared to those with lower levels. (*, all p<0.05). (D) The expression patterns for lung cancer patients and cancer-free controls show that miR-451 is detectable in all sample types with no significant difference between the patient and control groups. ns, not significant.

### RBC-derived miRNAs can be transferred to lung cancer cells predominantly via exosomal pathways

To explore whether RBC-derived miRNAs are transportable to lung cancer cells, we collected RBCs from two lung cancer patients, showing elevated miR-93-5p levels and those from two healthy controls with lower miR-93-5p levels. We then employed a Transwell system equipped with a porous membrane (0.4 μm) to divide the upper and lower chambers (Fig. S1). The system enabled direct contact between RBCs and lung cancer cells in the upper chamber, while cancer cells in the lower chamber were exposed to the same RBC-medium without direct RBC contact. Furthermore, cancer cells cultured with only regular medium served as control assays. After a 24-hour incubation, equal numbers of cancer cells were collected from both the upper and lower chambers, respectively, and RNA was isolated from each group. Equal amounts of RNA from the sets were subsequently analyzed to quantify miRNAs. Cancer cells incubated with RBCs and RBC medium from lung cancer patients in the up chamber showed miR-93-5p levels similar to those of cancer cells incubated with RBC medium alone in the low chamber (all p<0.01) (Fig. 2A). The miRNA levels in regular medium without RBCs and RBC medium were very low (Fig. 2A). Cancer cells cultured with RBCs from healthy controls exhibited low miR-93-5p expression in both upper and lower chambers, comparable to levels seen in cells grown in regular medium (all p<0.01) (Fig. 2A). miR-451, a housekeeping miRNA typical in RBCs, was consistently present at high levels in cancer cells in both chambers but absent in cells in regular medium only (p<0.01) (Fig. 2B). The uniform levels of both miR-93-5p and miR-451 in cancer cells between the upper chamber containing RBCs and their medium, and the lower chamber with only the RBC-medium, suggest that the medium itself, rather than direct cell contact, likely facilitates the transfer of RBC-derived miRNAs to cancer cells.

**Fig. 2.**
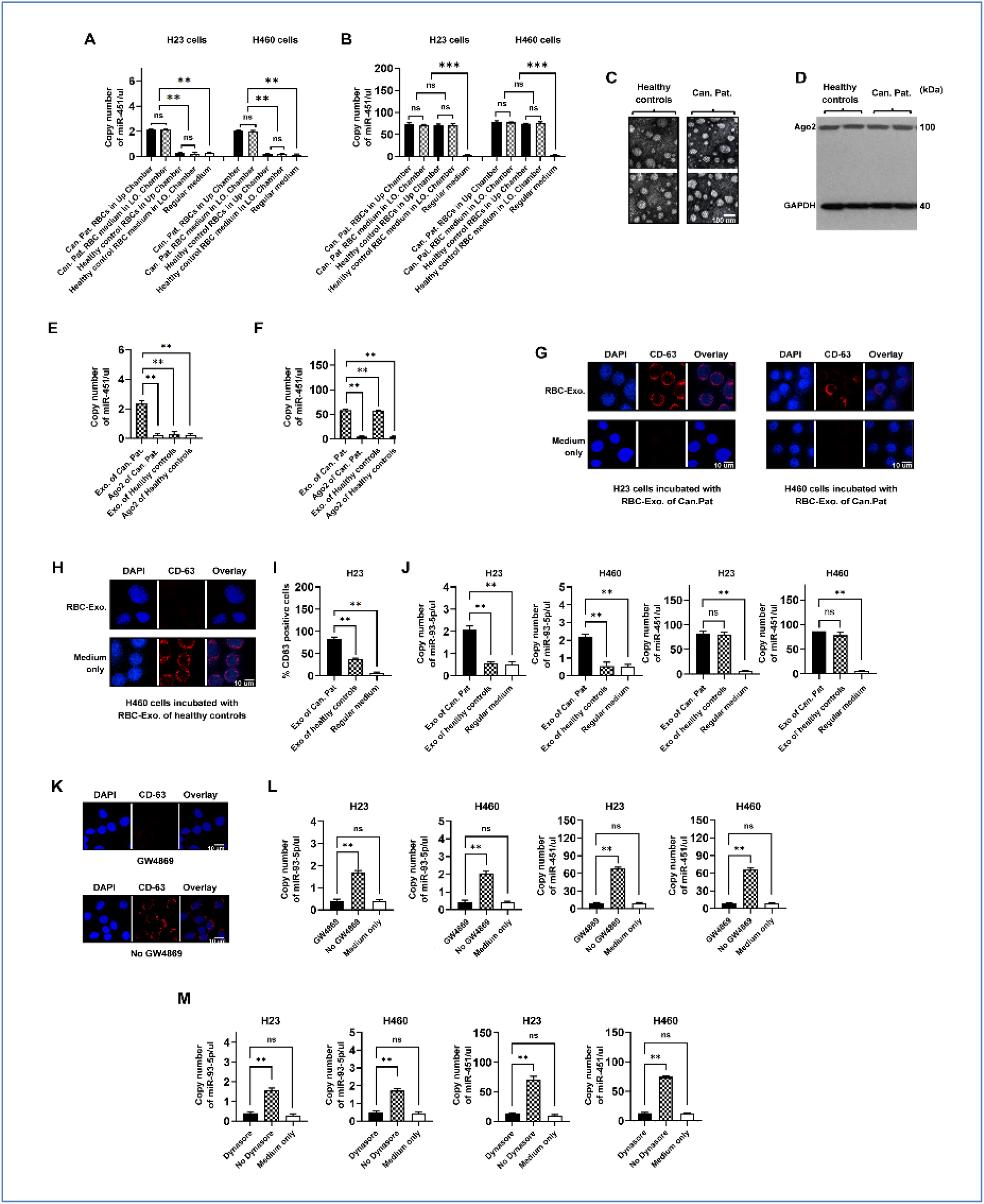
miRNA transfer from RBCs to lung cancer cells via exosomes (EXO.). (A) Relative levels of miR-93-5p in cancer cells after exposure to RBCs from cancer patients (Can. Pat.) and healthy controls, in upper chamber (Up chamber), lower chamber (LO. chamber), and regular medium as negative control. (B) Expression of miR-451 in cancer cells under the same experimental conditions as A. (C-D) Electron microscopy image and Western blot analysis demonstrating the isolation of exosomes and presence of Ago2 proteins of lung cancer patinets and healthy controls, respectively. (E) Quantification of miR-93-5p in isolated exosomes and Ago2-immunoprecipitated complexes, indicating significant presence in exosomes but not in Ago2 complexes. (F) Analysis of miR-451 levels in the same isolated fractions as in E. G) Fluorescent microscopy images showing the uptake of exosomes (red) marked with CD63 (exosome marker) in lung cancer cells with nuclei stained in blue. (H) Similar to G, showing uptake in cells incubated with exosomes from healthy controls. (I) The percentage of cancer cells positive for CD63 following incubation with exosomes sourced from both cancer patients and healthy controls. The exosomes released by RBCs of healthy controls exhibit CD63 labeling on cancer cells, indicating a baseline level of RBC exosomal activity. (J) Bar graphs displaying the levels of miR-93-5p and miR-451 in cancer cells after treatment with exosomes from cancer patients and healthy controls. (K) Effects of the exosome release inhibitor GW4869 on miRNA transfer from RBC-derived exosomes to cancer cells. Fluorescence images display nuclei stained with DAPI (blue) and CD63 immunostaining (red) in cancer cells. The up panel depicts cells treated with GW4869, showing an absence of red fluorescence, indicating reduced exosome uptake. The low panel shows untreated cells, displaying prominent red fluorescence. (L) The levels of miR-93-5p and miR-451 in cancer cells are lower after GW4869 treatment compared to untreated cells. (M) Effects of the exosome uptake inhibitor Dynasore on miRNA transfer. Cancer cells treated with Dynasore show lower levels of miR-93-5p and miR-451 compared to those without Dynasore treatment. ns, not significant, **, p<0.01, ***, p<0.001.

Given the lack of nucleus-based machinery in RBCs, it is proposed that exosomes and Argonaute 2 (Ago2) function as primary transporters of molecules crucial for facilitating communication from RBCs to other cells (*12, 16*). To explore the mechanism through which RBC-derived miR-93-5p is transferred from the RBC medium to lung cancer cells, we isolated exosomes and prepared immunoprecipitation products (IPs) with Ago2 from the RBC medium of two lung cancer patients and two healthy controls (Fig. 2C-D). RNA was purified from these exosomes and Ago2 IPs and then subjected to miRNA quantification. The exosomes exhibited significantly higher levels of miR-93-5p, whereas this miRNA was not detected in the Ago2 IPs from the same lung cancer patients (Fig. 2E). Both Ago2 IPs and exosomes from healthy controls displayed low miR-93-5p expression (Fig. 2E). Furthermore, while exosomes displayed significantly elevated levels of miR-451, this miRNA was undetectable in the IPs from both lung cancer patients and healthy controls (Fig. 2F). Altogether, the detection of miRNAs primarily in exosomes, rather than in Ago2 IPs, suggests that exosomes derived from RBCs likely serve as the principal transporters of the miRNAs, facilitating their conveyance to cancer cells.

To further determine whether exosomes facilitate the miRNA transfer, lung cancer cells were incubated with exosomes isolated from RBCs of cancer patients and healthy controls. The uptake of these exosomes by the lung cancer cells was verified through the identification of CD63, an established marker of exosomes (Fig. 2G). The release of exosomes from the RBCs of healthy controls resulted in CD63 labeling on cancer cells, marking a baseline level of RBC exosomal activity (Fig. 2H-I). The percentage of cells showing positive CD63 labeling was significantly lower compared to cells exposed to exosomes from the RBCs of cancer patients (p=0.001) (Fig. 2I). In line with this, cells treated with exosomes from cancer patients’ RBCs exhibited consistently higher levels of both miR-93-5p and miR-451 (Fig. 2J). However, cells exposed to exosomes from the RBCs of healthy controls exhibited elevated levels of miR-451 but not miR-93-5p (Fig. 2J).

We further employed the exosome release inhibitor (GW4869) to treat RBCs. Post-treatment, we harvested the conditioned medium from both GW4869-treated RBCs and RBCs without the treatment and introduced it to cancer cells for a 24-hour period. Exosomes were undetectable in cancer cells treated with GW4869 (Fig. 2K). Consequently, the absence of miR-93-5p and miR-451 in the cancer cells was observed (Fig. 2L), suggesting that inhibiting exosome release effectively blocks the transfer of miRNAs from RBCs to cancer cells. Moreover, we collected RBC-medium and co-incubated it with cancer cells, along with Dynasore, an inhibitor that blocks exosome uptake. After an incubation duration of 24 hours, there was no uptake of RBC-derived exosomes by the cancer cells, resulting in no subsequent increase in the levels of miR-93-5p and miR-451 within these cells (Fig. 2M). Conversely, cancer cells incubated with RBC-medium without the inhibitor were able to uptake RBC-derived exosomes, resulting in an increase in miR-93-5p and miR-451 levels (Fig. 2M). Collectively, our results highlight RBC-derived exosomes as a key vehicle for delivering miRNAs to cancer cells.

### The RBC-derived miR-93-5p promotes cell proliferation, invasiveness, and migration

We further investigated whether transferring RBC-miR-93-5p to lung cancer cells could enhance their tumorigenic properties. RBC-exosomes from four lung cancer patients and two healthy individuals were respectively incubated with cancer cells. The four lung cancer patients included two with high levels of miR-93-5p and two with low levels of miR-93-5p in their RBCs and exosomes. The cancer cells exposed to exosomes from the RBCs of the lung cancer patient with high levels of miR-93-5p exhibited elevated levels of miR-93-5p, which was accompanied by enhanced proliferation, invasion, and migration (Fig. 3A-E). Conversely, when cancer cells were incubated with exosomes from the RBCs of lung cancer patients with low levels of miR-93-5p and healthy donors, there was no increase in miR-93-5p levels, nor was there an effect on their malignant behaviors (Fig. 3A-E). Furthermore, cancer cells treated with RBC-exosomal miR-451 also did not exhibit increased malignant behaviors. Therefore, the transfer of miR-93-5p, a lung cancer associated RBC-miRNA, to cancer cells may enhance malignant properties, in contrast to the effect of transferring RBCs’ housekeeping miRNA, miR-451.

**Fig. 3.**
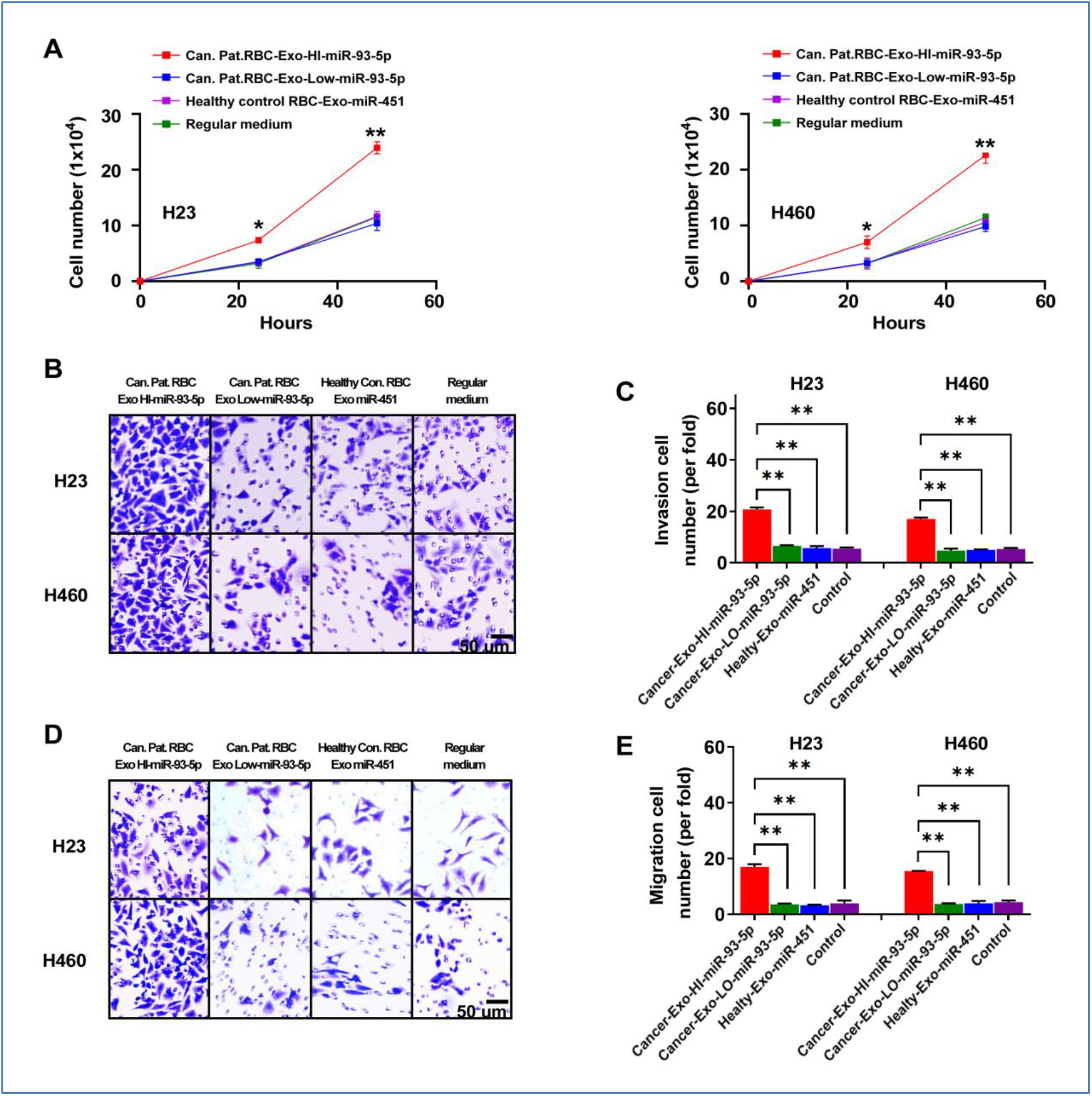
Effects of RBC-derived exosomal miR-93-5p on tumorigenicity of lung cancer cells. (A) Cancer cells, when incubated with RBC-exosomes from lung cancer patients with high levels of miR-95-3p, show increased proliferation compared to those incubated with RBC-exosomes from lung cancer patients with low miR-95-3p and healthy controls. (B) Invasion assays for cancer cells treated similarly demonstrate enhanced invasive capabilities with lung cancer patients RBC exosomal miR-95-3p. (C) Bars represent the invasive cells per field, confirming increased invasion with high miR-93-5p exosomes. (D) Migration assays for cancer cells treated with the same groups of exosomes. Cells exhibit increased migration capabilities when treated with exosomes from cancer patients with high miR-93-5p. (E) Quantification of migration assays shown in (D). Bars represent the migration cells per field, demonstrating enhanced migration with high miR-93-5p exosomal treatment. *, p<0.05., **, p<0.01.

### The transferred RBC-derived miR-93-5p targets the PTEN gene in cancer cells and promotes tumorigenesis

To unravel the mechanism through which RBC-derived miR-93-5p impacts the aggressive traits of cancer cells, we used prediction tools to identify genes potentially targeted by this miRNA. Bioinformatic analysis identified potential miR-93-5p binding sites on the 3’ Untranslated Region (UTR) of PTEN, a gene recognized for its tumor-suppressing roles, making it a focus for further investigation (Fig. 4A). A luciferase reporter assay was used to confirm the bioinformatic analysis by co-transfecting lung cancer cells with Wild-Type (WT) PTEN-3′UTR or its mutants (MUT) and a miR-93-5p mimic. Decreased luciferase activity in cells transfected with the WT construct suggests that miR-93-5p directly targets and binds to the mRNA, resulting in either its degradation or the inhibition of PTEN translation (Fig. 4B). Lung cancer cells exposed to RBC-derived exosomal miR-93-5p showed reduced PTEN protein expression compared with cells incubated with the control medium, corroborating the miRNA-mediated post-transcriptional suppression of PTEN (Fig. 4C). To elucidate whether the effect of miR-93-5p-enriched RBC-exosomes on cancer cells depended on PTEN expression, we introduced PTEN mimic into the cancer cells exposed to miR-93-5p. While miR-93-5p-rich RBC-exosomes notably enhanced the malignant properties of the cells, PTEN overexpression counteracted the effects of miR-93-5p on cell proliferation, migration, and invasion (Fig. 4D-G). Furthermore, we treated cancer cells with miR-93-5p-rich RBC-exosomes and then knockdown miR-93-5p in these cells using antisense oligonucleotide targeting microRNA-93-5p (ASO-miR-93-5p). Although the transfer of miR-93-5p to cancer cells enhanced their malignant behaviors, the use of ASO-miR-93-5p significantly decreased these effects (fig. S2). Taken together, these findings indicate that PTEN is a primary target of RBC-derived miR-93-5p, elucidating the cancer-promoting effects of the RBC-derived oncomiR on the malignant properties of cancer cells.

**Fig. 4.**
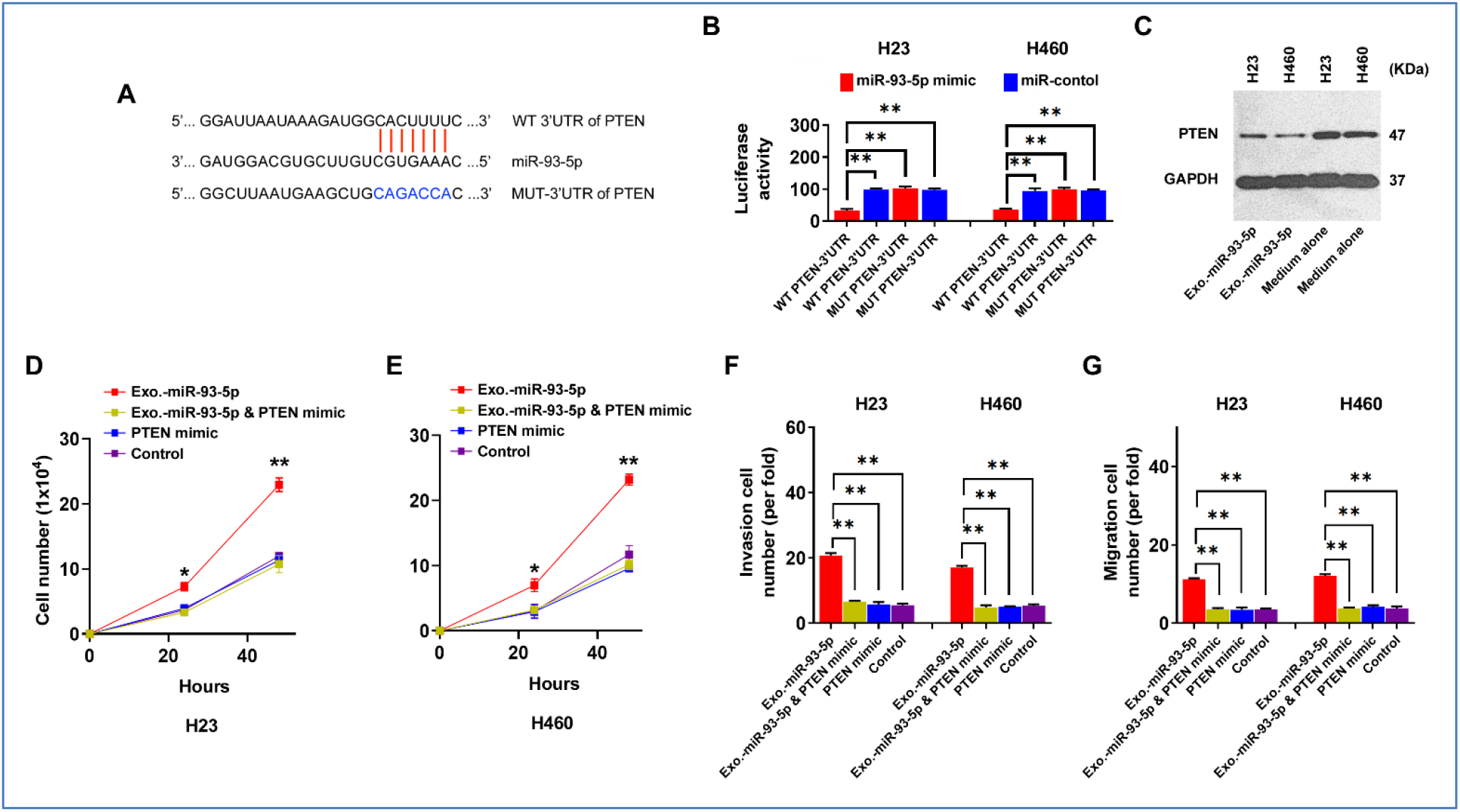
Impact of RBC-exosomal miR-93-5p on PTEN Regulation and Tumorigenicity in Lung Cancer Cells. (A) Sequence alignment highlighting the predicted binding sites of miR-93-5p on the 3’UTR of the PTEN gene, contrasting the wild type (WT) and mutant (MUT) versions. (B) Luciferase reporter assays displaying reduced activity in lung cancer cells co-transfected with WT PTEN 3’UTR, suggesting direct interaction and targeting. (C) Western blot analysis showing that lung cancer cells treated with RBC-exosomal miR-93-5p exhibit decreased PTEN protein levels compared to those treated with regular medium only. (D-E) The effect of PTEN overexpression on cell proliferation in cancer cells treated with miR-93-5p-rich exosomes, indicating a suppressive effect on proliferation due to PTEN. (F-G) The impact of PTEN on invasion and migration of cancer cells incubated with miR-93-5p-rich exosomes. *, p<0.05., **, p<0.01.

### RBC-derived miR-93-5p promotes growth of lung tumor in xenograft animal models

The mice were randomly divided into two groups: Group 1 received subcutaneous injections of H460-Luc cells treated with RBC-derived exosomes expressing low levels of miR-93-5p from a healthy individual. Conversely, Group 2 was injected with H460-Luc cells treated with RBC-derived exosomes that exhibit high expression of miR-93-5p from lung cancer patients. By the end of the fourth week, tumors derived from the Group 2 were notably larger, with an average volume of 88.39±14.26 mm ^3^, in contrast to the tumors in group 1, which averaged 25.75±15.48mm^3^ (p=0.049) (Fig. 5A) (fig. S3). These tumors demonstrated reduced PTEN expression and elevated KI-67 levels when compared to tumors from the control group (all p<0.05) (Fig. 5B). The results from the xenograft mouse model further support the crucial role of RBC-derived miR-93-5p in promoting lung cancer development by targeting PTEN.

**Fig. 5.**
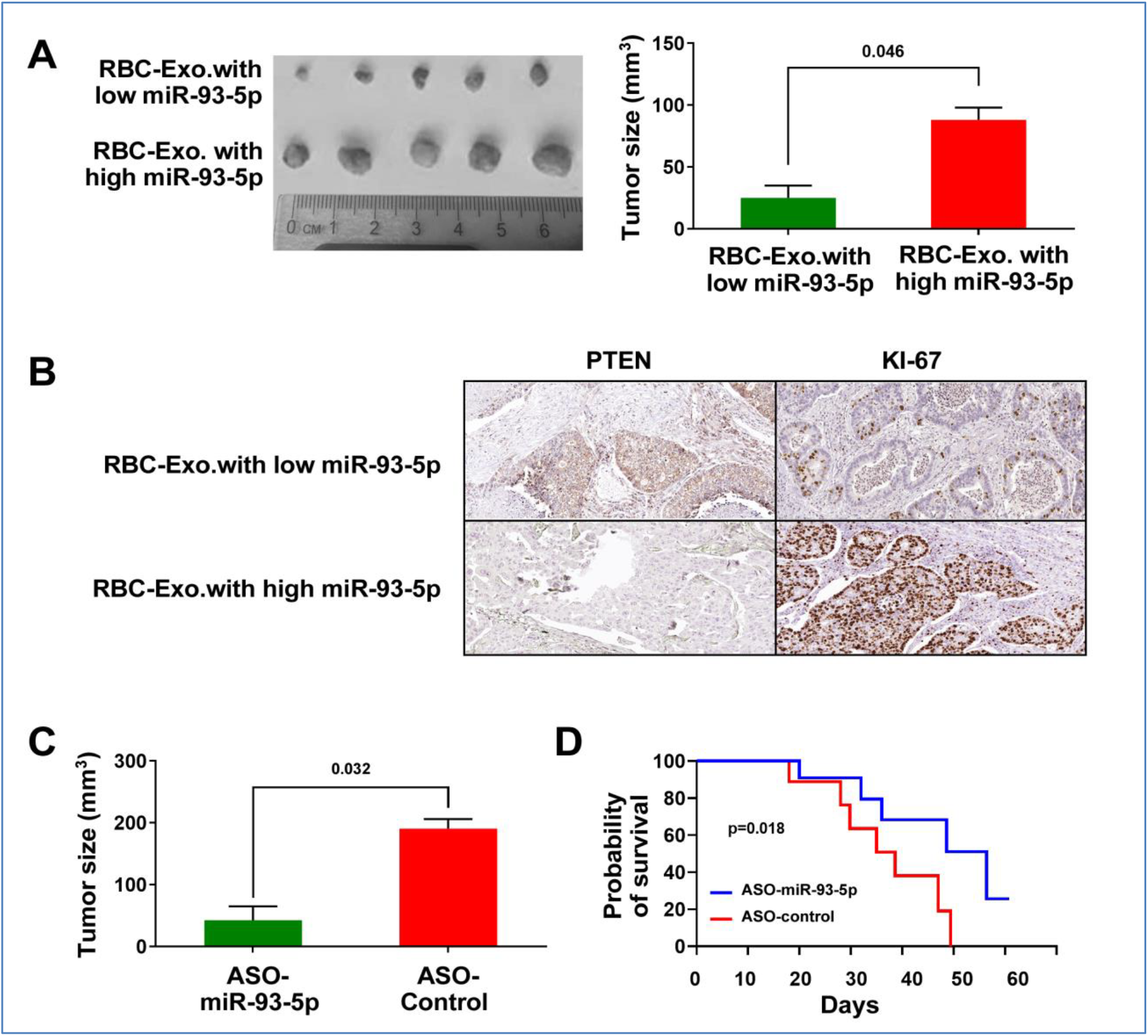
Effects of RBC-Derived miR-93-5p on Tumor Development in Lung Cancer Xenograft Mouse Models and the Therapeutic Potential of RBC-Derived miR-93-5p Inhibition. (A) On the left, extracted tumors at the end of week 4 from each group are displayed, demonstrating visibly larger tumors in the high miR-93-5p group. The graph on the right quantifies this difference, showing a significant increase in tumor volume in the high miR-93-5p group (p=0.046). (B) Immunohistochemical staining for PTEN (tumor suppressor) and KI-67 (proliferation marker) in tumors from mice injected with low vs. high miR-93-5p RBC-derived exosomes. The images were shown at 20x magnification. Tumors with high miR-93-5p show reduced PTEN expression and increased KI-67 levels. (C) Mice with tumors derived from cells treated with high miR-93-5p exosomes were given direct tumor injections with ASO-miR-93-5p or ASO-control. The graph indicates a significant reduction in tumor volume in the ASO-miR-93-5p treatment group (p=0.032). (D) Kaplan-Meier survival curves for mice treated with ASO-miR-93-5p (blue line) versus ASO-control (red line). Mice receiving the miR-93-5p-targeting ASOs demonstrated significantly extended survival (p=0.018).

### Effective antitumor effects ofASO-miR-93-5p in vivo

To explore the impact of inhibiting RBC-derived miR-93-5p on tumor growth in vivo, mice were injected with H460-Luc cells treated with RBC-derived exosomes with high expression of miR-93-5p from lung cancer patients. Within 10 days, tumors had formed in all the mice injected with cancer cells. Subsequently, these mice were randomly divided into two groups. Group 1 received direct injections into the tumors with ASO-miR-93-5p, while Group 2 received a control ASO, administered three times per week for three weeks. Tumors treated with ASO-miR-93-5p exhibited a significantly larger reduction in volume compared to the control group (190.52±21.92 mm^3^ vs. 42.39±31.82 mm^3^, p=0.032) (Fig. 5C). Furthermore, mice receiving ASO-miR-93-5p exhibited a longer survival period than those treated with control ASO (p=0.018) (Fig. 5D). Thus, miR-93-5p inhibition could effectively reduce tumor growth in vivo, highlighting its potential as a therapeutic strategy.

## Discussion

Lung cancer is the leading cause of cancer-related mortality worldwide among both men and women. Understanding the mechanisms of tumorigenesis is critical for developing effective prevention, diagnosis, and treatment strategies. Our study demonstrates that elevated levels of miR-93-5p in RBCs and their derived exosomes correlate with reduced survival rates. We further reveal a previously unrecognized role of RBCs in lung tumorigenesis, where they promote cancer progression through the transfer of oncogenic miR-93-5p via exosomes. These findings deepen our understanding of lung cancer pathogenesis and may offer novel prognostic and therapeutic strategies, emphasizing the significant role of RBCs in tumor progression.

Mature RBCs, which constitute about 45% of blood volume, carry a diverse array of miRNAs, despite their enucleated state. (*17*). However, the roles of RBCs and RBC-miRNAs in tumorigenesis remain poorly understood. Since RBCs lack a nucleus, the Ago2 complex and exosomes have been proposed as the main vehicles for transferring RBC-derived molecules in communication from RBCs to other cells (*12, 16*). Our current research indicates that elevated levels of miR-93-5p are found primarily in exosomes, not in Ago2 complex. Furthermore, inhibiting exosome release from RBCs or blocking their uptake by cancer cells can effectively halt the transfer of miRNAs from RBCs to cancer cells. In line with these findings, exosomes derived from RBCs are capable of significantly increasing miR-93-5p levels in lung cancer cells. Our findings emphasize how RBCs contribute to carcinogenesis through exosomal pathways that transfer miRNAs to lung cancer cells, thereby modulating tumorigenic properties and highlighting the complex dynamics of RBC-to-cell communication in cancer biology.

miR-93-5p is recognized as an oncomiR in tumorigenesis (*18*). This miRNA can activate the PI3K/Akt signaling pathway which further promotes tumor cell malignancy(*19*). Additionally, miR-93-5p can inhibit NEDD4L and ZNRF3, subsequently affects the epithelial-mesenchymal transition and Wnt signaling pathways (*20*). Our bioinformatics analysis and luciferase reporter assays revealed that miR-93-5p, derived from RBCs, upon transfer to cancer cells, can target PTEN. Furthermore, targeting PTEN with miR-93-5p in lung cancer cells leads to an increase in malignant characteristics, including cell proliferation, invasion, and migration. These effects were reduced when PTEN was overexpressed or when miR-93-5p was inhibited, confirming the specificity of miR-93-5p’s action through exosomal delivery and its pivotal role in modulating PTEN. Our in vivo animal studies further confirm that miR-93-5p derived from RBCs facilitates the growth of lung tumors. Conversely, effective antitumor effects have been observed with miR-93-5p-targeting ASOs in vivo. While RBC-derived miR-451 can also be transferred to cancer cells via exosomes, it does not affect their malignancy due to its housekeeping role in RBCs. Furthermore, miR-451 is downregulated in tumors and may act as a tumor suppressor(*21*), contrasting with the oncogenic miR-93-5p, whose elevation does not influence cancer malignancy similarly. Therefore, our findings collectively highlight the critical role of RBC-derived exosomal miR-93-5p, an oncomiR, in lung cancer progression through the downregulation of PTEN, underscoring its potential as a therapeutic target in cancer treatment.

Future research should focus on tracing the origin and packaging process of oncomiRNAs, such as miR-93-5p, within RBCs, derived from their precursor cells, to unveil its role in miRNA-mediated oncogenic signaling. Moreover, investigating the therapeutic potential of targeting pathways involving RBC-derived miR-93-5p could offer innovative strategies to interrupt oncogenic signaling in lung cancer. Additionally, utilizing the longevity and compatibility of RBCs to transport ASOs or nucleic acid-based molecules targeting miR-93-5p directly to tumors may emerge as a promising cancer treatment approach.

In conclusion, our study discovers a novel biological function of RBCs in tumorigenesis. We reveal a mechanism by which RBCs promote the progression of lung cancer through the transfer of oncomirs via the exosomal pathway. These findings have the potential to open new avenues for the development of diagnostic and therapeutic strategies against lung cancer.

## Material and Methods

### Patients and specimens

Our study was approved by the Institutional Review Board of University of Maryland Baltimore. We recruited non-small lung cancer (NSCLC) patients and cancer-free smokers based on criteria complemented by demographic, radiological, and clinical data from their medical records [22]. Blood samples were collected from the participants and plasma was separated from blood samples by following established clinical protocols [23-25]. 58 NSCLC patients and 45 cancer-free smokers were enrolled (Supplementary Table 1). Among the NSCLC patients, stages I to IV were represented, with 33 cases of adenocarcinoma and 25 of squamous cell carcinoma. We had complete medical records and follow-up data for all 58 patients with NSCLC. Survival time was calculated from the date of diagnosis to the date of last follow-up or death.

### Cell culture

Human NSCLC cell lines H23 and H460 were obtained from the American Type Culture Collection (ATCC, Manassas, VA). These cell lines were meticulously cultured in accordance with the manufacturer’s instructions, ensuring optimal growth conditions and the maintenance of cell line integrity for experimental use.

### Droplet digital PCR (ddPCR)

ddPCR was employed for quantifying miRNA copy numbers following RNA isolation, as previously detailed(*13*) . RNA underwent reverse transcription employing the TaqMan miRNA RT Kit (Thermo Fisher Scientific., Waltham, MA). ddPCR reactions were prepared, including a mix of cDNA, Supermix, and Taqman primer/probe mix, and were processed in the QX100 Droplet Generator (Bio-Rad Laboratories, Hercules, CA). This process yielded over 10,000 droplets per well. Following PCR amplification, droplet analysis was performed using a fluorescence detector (Bio-Rad Laboratorie), enabling the accurate detection of miRNA levels. The concentration of target genes was determined utilizing Poisson’s distribution.

### Isolation of RBCs and RBC-exosomes

Fresh blood collected in Ethylenediaminetetraacetic acid tubes (BD, Franklin Lakes, NJ) was centrifuged at 1,500 x g for 10 minutes to separate the plasma and buffy coat from the RBCs, as previously described (*22*). The RBC pellet was then resuspended in a hypotonic buffer (10 mM Tris-HCl at pH 7.4) (Sigma-Aldrich, St. Louis, MO) and incubated for 30 minutes. The sample was then centrifuged at 300 x g for 10 minutes. We collected the supernatant and further centrifuged it at 16,500 x g for 20 minutes. We performed ultracentrifugation of the supernatant at 100,000 x g for 70 minutes at 4°C to pellet the exosomes. The exosome pellet was resuspended in PBS (Sigma-Aldrich).

### Electron microscopy for the analysis of isolated exosomes

The exosomal samples were fixed in 2.5% glutaraldehyde in 0.1 M cacodylate buffer (Sigma-Aldrich). The samples then underwent a wash in cacodylate buffer (Sigma-Aldrich) and were then dehydrated through a graded ethanol series (Sigma-Aldrich). The exosome suspension was placed on microscopy grid (Ted Pella, Inc., Redding, CA) for 20 minutes. The sample was negatively stained with 2% uranyl acetate (Sigma-Aldrich) for one minute. The grid was loaded into an Transmission Electron Microscopes (TEM) (Thermo Fisher Scientific) to capture images for size, shape, and aggregation of exosomes.

### Immunofluorescent staining of exosomes with CD63

Exosomes were affixed onto slides using paraformaldehyde (Sigma-Aldrich). The samples were treated with 0.1% Triton X-100 (Sigma-Aldrich). Non-specific binding was blocked by incubating the slides with bovine serum albumin (Sigma-Aldrich) for one hour. The slides were then incubated with a primary antibody against CD63 (Abcam, Cambridge, MA), diluted in blocking buffer (Sigma-Aldrich), overnight at 4°C. After washing the slides three times with PBS (Sigma-Aldrich), the slides were incubated with a fluorescently labeled secondary antibody (conjugated with Alexa Fluor) (Abcam). Nuclei were stained using DAPI (Sigma-Aldrich). The slides were examined under a fluorescence microscope (Zeiss, Germany) to visualize the stained CD63 and nuclei.

### Investigating RBC-derived miRNA impact on lung cancer via Transwell System

RBCs were collected from lung cancer patients with elevated miR-93-5p and healthy individuals with lower levels of miR-93-5p. A Transwell insert (0.4 μm pore size) (Corning Inc. Corning, NY) was used to created upper and lower chambers, with RBCs placed in the upper chamber. Care ensured cancer cells in the lower chamber were exposed to the same medium without RBC contact. After a 24-hour incubation, equal cancer cell numbers were collected from both chambers. RNA was isolated and analyzed using ddPCR to quantify miRNAs.

### Exosome release and uptake inhibition

1×10^5^ cancer cells were treated with 10 ng/µl of GW4869 (MilliporeSigma, Billerica, MA), an exosome release inhibitor, for 24 hours. Conditioned medium from both GW4869-treated and untreated control RBCs was collected. One ml of conditioned medium from both GW4869-treated and control RBCs was introduced to 1×10^5^ cancer cells and incubated for 24 hours. Additionally, 1 ml of RBC-medium was collected and co-incubated with 1×10^5^ cancer cells, followed by the addition of 5 ng/µl of Dynasore (MilliporeSigma), an inhibitor of exosome uptake, to the co-incubation mixture for 24 hours.

Exosome presence in cancer cells was assessed as described above, utilizing CD-63 for detection. miRNA levels were detected using ddPCR.

### Immunoprecipitation (IP) products with Ago2

Samples were centrifuged at 10,000 x g for 10 minutes to remove cellular debris as previously described(*23*). The supernatant was treated with a protein extraction buffer containing RNase inhibitors and detergents (Sigma-Aldrich), then incubated with protein A/G magnetic beads (Sigma-Aldrich) at 4°C for an hour. After magnetic separation of the beads, an Ago2-specific antibody (Abcam) was added to the supernatant and incubated overnight at 4°C. Protein A/G magnetic beads (Sigma-Aldrich) were then used to capture the antibody-antigen complexes by incubating for 2 hours at 4°C. After washing the beads 3 times with cold buffer (Sigma-Aldrich), the complexes were eluted and analyzed by Western blotting to verify the presence of Ago2.

### Cell transfection

We utilized synthetic constructs provided by Integrated DNA Technologies (IDT, Coralville, IA), including miR-93-5p mimics and miR-93-5p antisense oligoribonucleotide (ASO), along with corresponding nonsense controls (NC). The sequences were as follows: miR-93-5p mimic, Forward 5′-CAAAGUGCUGUUCGUGCAGGUAG-3′ and Reverse 5′-CACCUGCACGAACAGCACUUUGU-3′; NC mimic, Forward 5′-UUCUCCGAACGUGUCACGUTT-3′ and Reverse 5′-ACGUGACACGUUCGGAGAATT-3′; miR-93-5p ASO, 5′-CACUUAUCAGGUUGUAUUAUAA-3′; and NC ASO, 5′-CAGUACUUUUGUGUAGUACAA-3′ (*24*). Lipofectamine RNAiMAX (Life Technologies, Gaithersburg, MD) was employed to ensure uniform transfection conditions across all cancer cell line experiments.

### MTT Assay (3-(4,5-Dimethylthiazol-2-yl)-2,5-diphenyltetrazolium bromide)

Cells were seeded at a density of 5,000 cells per well in a 96-well plate and incubated at 37°C with 5% CO2, as outlined in previous study (*25*). 100 μL of fresh medium containing 0.5 mg/mL MTT (Sigma-Aldrich) was added to each well, and the cells were incubated at 37°C. Following this period, the MTT solution was removed, and 100 μL of Dimethyl sulfoxide (Sigma-Aldrich) was added to solubilize the formazan crystals. Absorbance at 570 nm was measured using a microplate reader. Cell viability was assessed by calculating and normalizing the average absorbance for each treatment group against the control group.

### Cell invasion assay

We seed cells into the upper chamber of a Transwell device (Corning Inc., Corning, NY), which is coated with Matrigel (BD Biosciences, Franklin Lakes, NJ). After incubation in the chambers, cells migrating to the lower chamber are fixed with methanol, stained with crystal violet, and then counted to assess their invasive capabilities.

### Cell migration assays

We used the Transwell assay (AMSBIO LLC, Cambridge, MA) to determine cell migration (*25*). The cells that had migrated to the lower surface of the membrane were fixed with formalin and stained with crystal violet (Sigma-Aldrich). The migrating cells were examined microscopically and determined by counting the migrating/invasive cells in 5 randomly selected fields using an Olympus BX41 microscope (Olympus Corporation).

### Dual luciferase reporter assay

We inserted PTEN 3′UTRs with predicted binding sites for miR-93-5p into pmirGLO vectors (Promega Corporation, Madison, WI). These constructs, along with miR-93-5p mimics and inhibitors, were transfected into lung cancer cells to study miR-93-5p’s regulatory effects on PTEN using the dual-luciferase reporter assay (Thermo Fisher Scientific).

### Western blot analysis

We separated 30 µg of protein samples using SDS-PAGE (Sigma-Aldrich), then transferred them onto PVDF membranes (MilliporeSigma). The membranes were incubated with primary antibodies (Abcam), followed by secondary antibodies. Detection was performed using an enhanced chemiluminescence reagent (MilliporeSigma).

### Evaluation of miR-93-5p’s oncogenic role in a human lung cancer xenograft model

In accordance with an approved animal study protocol from the University of Maryland Baltimore, we utilized six-week-old female immunodeficient NOD. Athymic Swiss nu/nu/Ncr nu (nu/nu) mice sourced from The Jackson Laboratory (Bar Harbor, ME). To assess the in vivo oncogenic potential of RBC-exosomal miR-93-5p, the mice were divided into two groups, each comprising five animals. The experimental group received subcutaneous injections of 1 × 105 NCI-H460-luc cells (TD2 Precision Oncology, Scottsdale, AZ) pre-incubated with 50 ng/μl of exosomes derived from healthy donors. Conversely, the control group received the same number of cells pre-incubated with exosomes from lung cancer patients with elevated miR-93-5p levels, as confirmed by ddPCR. Tumor progression was monitored weekly using the IVIS 200 Imaging System (Xenogen, Alameda, CA). After four weeks, the mice were humanely euthanized under deep anesthesia induced by pentobarbital (Sigma-Aldrich), and tumor dimensions were quantified using the standard volumetric formula. Histological and molecular analyses were conducted on the harvested tissue samples. To further validate the therapeutic targeting of miR-93-5p, twenty additional mice were subcutaneously injected with A549-Luc cells (CCL-185-LUC2™, ATCC) and treated with either a ASO-control or a ASO-miR-93-5p (AstraZeneca, Wilmington, DE). Tumor growth was monitored weekly using the IVIS system. After four weeks, tissue analyses were performed to elucidate molecular changes, and survival outcomes were monitored to assess the impact of miR-93-5p modulation on overall survival.

### Immunohistochemical analysis of tumor tissues

Tumor tissues were fixed, paraffin-embedded, and sectioned for analysis. After antigen retrieval, the sections were incubated with primary antibodies (Abcam), followed by a secondary antibody. Chromogen development and counterstaining were then performed, with observations made under a microscope (Zeiss). Quantitative analysis involved selecting five representative regions, counting positively stained cells, and averaging the counts per 100 cells. Staining intensity was graded on a scale from 0 (0%-5% positive cells) to 3 (over 50% positive cells). Manual counts were done using AnalySIS Pro software (Olympus), with independent repeats by two scientists to ensure accuracy. All analyses were blinded to minimize bias.

### Bioinformatics analysis for gene interaction

The interaction between lncRNA and miRNA was predicted using bioinformatics tools: starBase 3.0 and miRDB. Putative targets of miRNA were identified using TargetScan and miRDB.

### Statistical analysis

We used t-test to determine the statistical significance of molecular changes and clinical factors between cases and controls. We used Pearson’s correlation analysis to explore relationships between molecular alterations and various clinical parameters, including histopathologic characteristics of lung tumors, clinical stages, and patient outcomes. Survival distributions of lung cancer patients were estimated using the Kaplan-Meier method, and differences between survival curves were analyzed using the log-rank test. For animal survival data, the Kaplan-Meier method was also utilized to compute survival estimates, with the log-rank test applied to compare survival across different experimental groups.

## Data Availability

All data produced in the present work are contained in the manuscript

## Author contributions

Conceptualization: FJ, Methodology: JL, PD, VH, AS. Funding: FJ. Supervision: FJ. Writing – JL and FJ.

## Data and materials availability

All data are available in the main text or the supplementary materials.

## Supplementary Materials Figs. S1 to S3

**Fig. S1.**
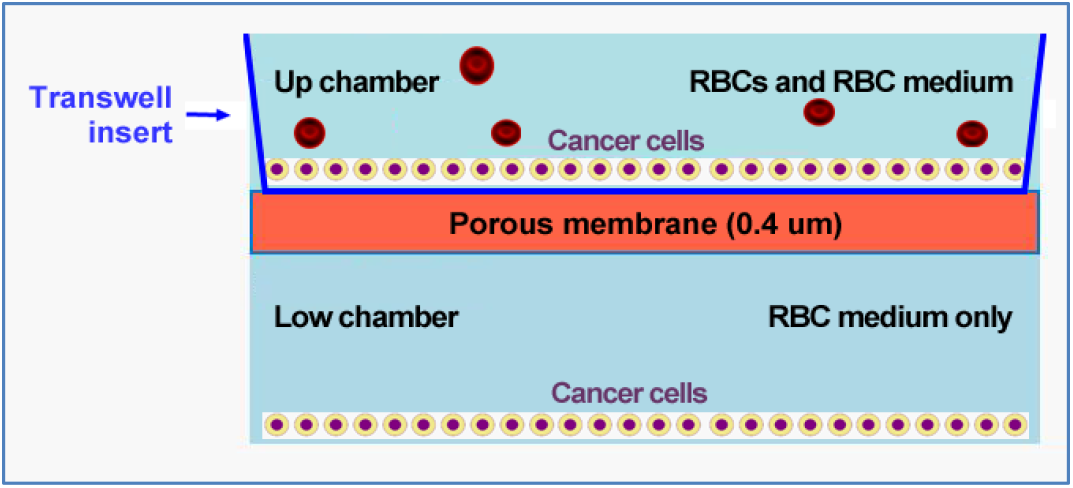
The diagram depicts a dual-chamber Transwell system utilized to study the transport of RBC-derived miRNAs to lung cancer cells. The system is divided by a porous membrane with a pore size of 0.4 micrometers, facilitating the selective transfer of RBC medium between the chambers. In the upper chamber, lung cancer cells are in direct contact with a mixture of RBCs and RBC medium. In contrast, the lower chamber contains another layer of lung cancer cells that are exposed only to the RBC medium, without direct RBC contact.

**Fig. S2.**
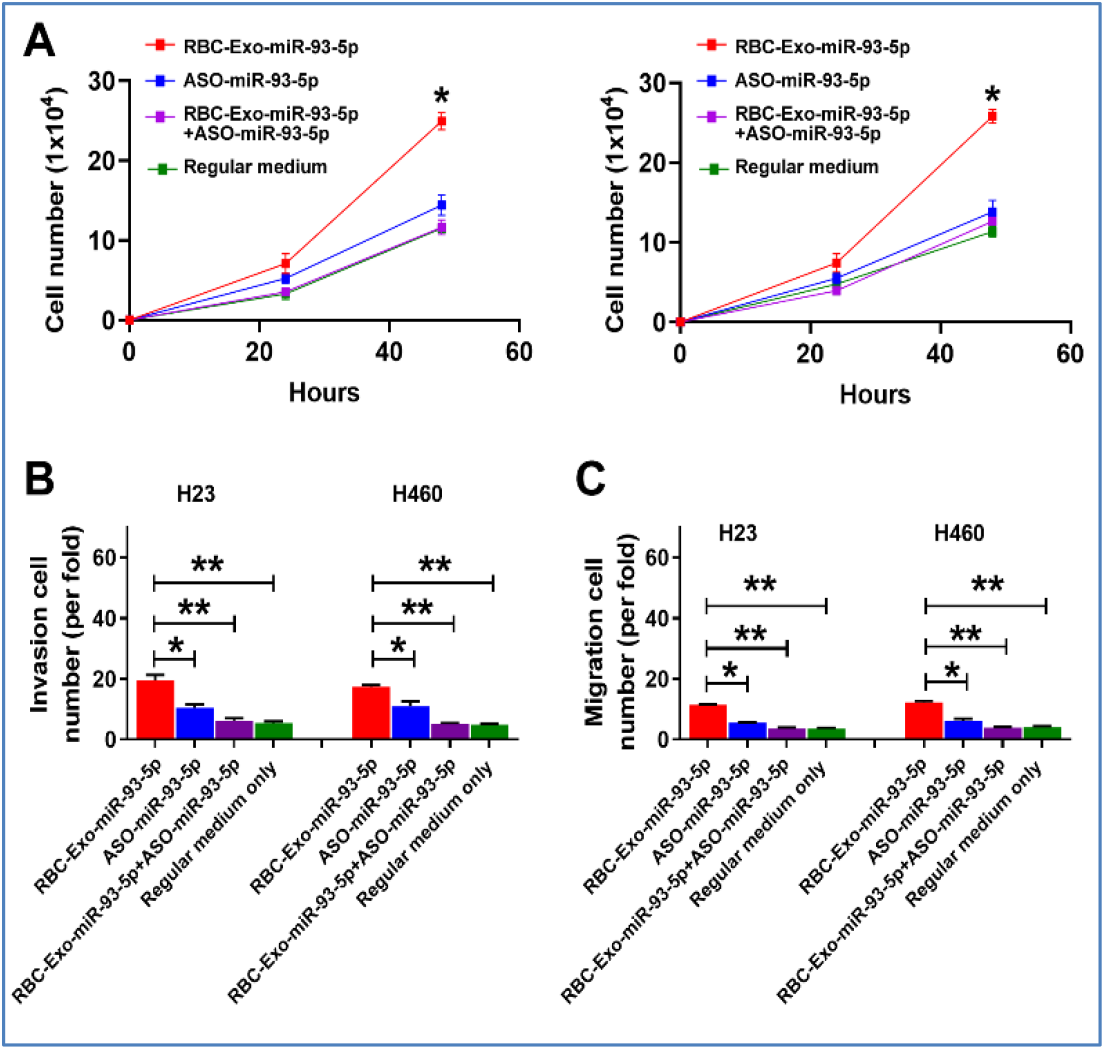
Effects of miR-93-5p-rich RBC-exosomes and ASO-miR-93-5p on tumorigenicity of cancer cells. (A) Cells treated with cancer-derived exosomes enriched in miR-93-5p (Cancer-Exo-miR-93-5p) have increased proliferation over time, compared to the negative control. The ASO-miR-93-5p treatment reduces cell numbers compared to cells treated with miR-93-5p-rich exosomes, approaching the levels of regular medium. B) Invasion assay results for cancers cells treated with RBC-exosomes rich in miR-93-5p from lung cancer patients, ASO-miR-93-5p, and control conditions, represented as the fold change relative to the negative control. In both cell lines, RBC-exosomes rich in miR-93-5p from lung cancer patients enhance invasion capabilities, which are significantly decreased by ASO-miR-93-5p treatment. C) Migration assay results for cancer cells under the same treatment conditions as in (B), showing a similar trend where miR-93-5p increases cell migration, and this effect is mitigated by ASO-miR-93-5p. Error bars indicate standard deviation. *, p<0.05, **, p<0.01.

**Fig. S3.**
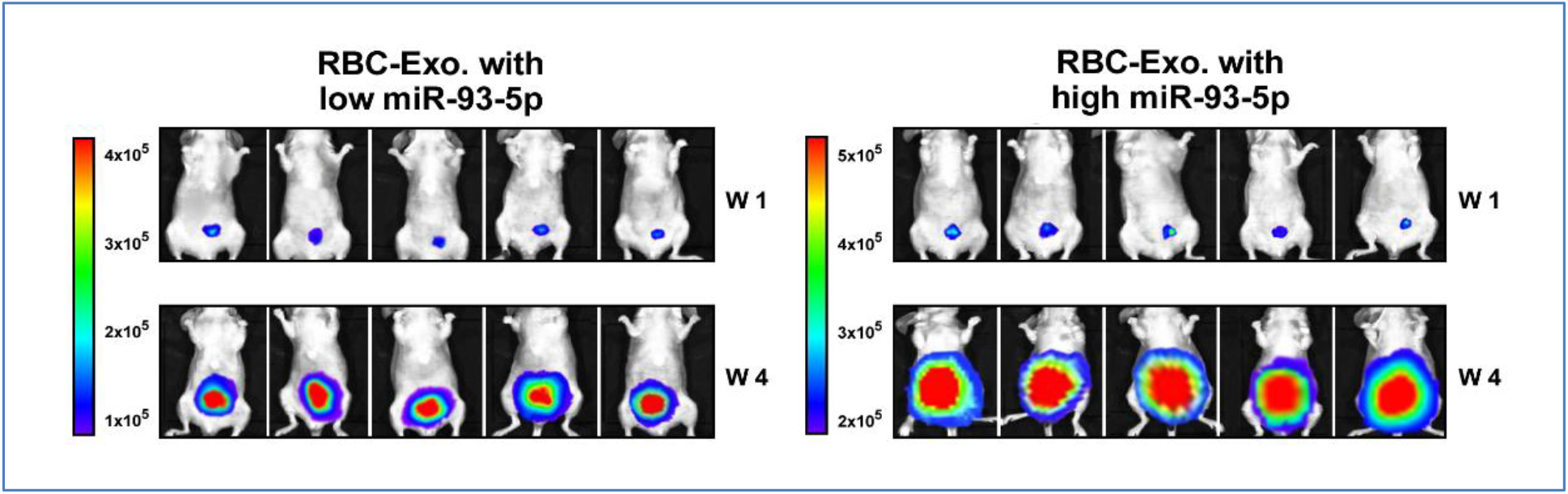
Mice were injected with H460-Luc cells pre-treated with RBC-derived exosomes containing either low (left) or high (right) levels of miR-93-5p. Bioluminescence Imaging at week 1 and week 4 shows increased tumor growth in mice receiving cells with high miR-93-5p exosomes.

## Tables S1 to S2

**Table S1.** Characteristics of NSCLC patients and cancer-free smokers.

| Characteristics | NSCLC cases (n = 58) | Controls (n = 45) |
| --- | --- | --- |
| Age | 68.37 (SD 11.58) | 66.23 (SD 11.19) |
| Sex |  |  |
| Female | 20 | 16 |
| Male | 38 | 29 |
| Race |  |  |
| African Americans | 21 | 16 |
| White Americans | 37 | 28 |
| Smoking pack-years (median) | 32 | 27 |
| Stage |  |  |
| Stage I | 17 |  |
| Stage II | 32 |  |
| Stage III-IV | 9 |  |
| Histological type |  |  |
| Adenocarcinoma | 33 |  |
| Squamous cell carcinoma | 25 |  |

Abbreviations: NSCLC, non-small cell lung cancer. SD, standard deviation.

**Table S2.** Associations between RBC-miR-93-5p in RBCs and exosomes and clinical and demographic data, analyzed using Pearson’s correlation coefficients.

|  | RBC-miR-93-5p | Exosomal miR-93-5p | Age | Gender | Pack-smoking years | Types of cancer | Stage |
| --- | --- | --- | --- | --- | --- | --- | --- |
| RBC-miR-93-5p | 1 | 0.023 | 0.242 | 0.071 | 0.019 | 0.262 | 0.048 |
| Exosomal miR-93-5p |  | 1 | 0.155 | 0.239 | 0.025 | -0.159 | 0.029 |
| Age |  |  | 1 | -0.304 | 0.115 | 0.293 | 0.094 |
| Gender |  |  |  | 1 | 0.327 | 0.218 | 0.125 |
| Pack-smoking years |  |  |  |  | 1 | 0.143 | 0.203 |
| Types of cancer |  |  |  |  |  | 1 | 0.137 |
| Stage |  |  |  |  |  |  | 1 |

